# CD19/CD20 Bispecific Chimeric Antigen Receptor (CAR) in Naïve/Memory T Cells for the Treatment of Relapsed or Refractory Non-Hodgkin Lymphoma

**DOI:** 10.1101/2022.09.13.22279873

**Authors:** Sarah M. Larson, Christopher M. Walthers, Brenda Ji, Sanaz N. Ghafouri, Jacob Naparstek, Jacqueline Trent, Jia Ming Chen, Mobina Roshandell, Caitlin Harris, Mobina Khericha Gandhi, Thomas Schweppe, Beata Berent-Maoz, Stanley B. Gosliner, Amr Almaktari, Martin Allen-Auerbach, Jonathan Said, Karla Nawaly, Monica Mead, Sven De Vos, Patricia Young, Caspian Oliai, Gary J. Schiller, John M. Timmerman, Antoni Ribas, Yvonne Y. Chen

## Abstract

To address antigen escape and loss of T-cell functionality, we report a phase-1 clinical trial (NCT04007029) evaluating autologous naïve and memory T (T_N/MEM_) cells engineered to express a bispecific anti-CD19/CD20 CAR (CART19/20) for patients with relapsed/refractory NHL, with safety as the primary end point. Ten patients were treated with 36–165 × 10^6^ CART19/20 cells. No patient experienced neurotoxicity of any grade, or over grade-1 cytokine release syndrome. One case of dose-limiting toxicity (persistent cytopenia) was observed. Nine of ten patients achieved objective response (90% ORR), with seven achieving complete remission (70% CR rate). One patient relapsed after 18 months in CR, but returned to CR after receiving a second dose of CART19/20 cells. Median progression-free survival and overall survival were not reached with a 17-month median follow-up. In conclusion, CART19/20 T_N/MEM_ cells are safe and effective in patients with relapsed/refractory NHL, with durable responses achieved at low dosage levels.

## INTRODUCTION

Effective chimeric antigen receptor (CAR)-T cell therapy exerts a strong selective pressure against cancer cells that express the CAR-targeted antigen, and downregulation or loss of expression is the natural escape route for target antigens that are not critical to cell survival. Accurate quantification of relapse rate attributable to CD19 antigen escape is complicated by lack of tissue acquisition following relapse, and reported CD19-negative relapse frequencies range from 27% to 100% of relapsed cases among patients with leukemia and lymphoma (1-5). The frequency of cases with CD19-negative relapse demonstrates the susceptibility of CD19 to antigen loss, and points to the identification of alternative target antigens that are more resistant to gene-expression downregulation as a potential remedy.

To address the problem of CD19 antigen escape, we developed a CD19/CD20 bispecific CAR-T cell therapy, and previously demonstrated its ability to eradicate B-cell lymphoma with heterogenous CD19 expression and prevent relapse in mouse models of human lymphoma (6,7). CD19/CD20 bispecific CAR-T cells outperformed single-input CD19 CAR-T cells in achieving long-term, progression-free survival in a lymphoma xenograft model (6,7). CD20, like CD19, is pan–B-cell marker, and the first-line therapy for B-cell malignancies typically includes an anti-CD20 antibody such as rituximab (8). In fact, rituximab is commonly included in each subsequent line of chemotherapy administered to patients with NHL, yet CD20 antigen loss is a low-frequency event despite repeated cycles of CD20-targeted therapies (9), suggesting CD20 may be a suitable CAR target with low propensity for antigen escape. However, the clinical outcomes of CD20 CAR-T cell therapy have been uneven to date (10-13), resulting in more limited clinical advancement compared to CD19 CAR-T cell therapy. We hypothesized that simultaneously targeting CD19 and CD20 would both enable high initial response rate and increased resistance to antigen escape. Importantly, dual targeting of CD19 and CD20 would not increase on-target, off-tumor toxicity compared to either CD19 or CD20 single-input CAR-T cell therapy because both CD19 and CD20 are B-cell–specific markers, thus limiting the off-tumor toxicity to healthy B cells whose aplasia is a clinically manageable condition (14).

Multiple trials have reported association of CD19-positive relapses with T-cell exhaustion and lack of CAR-T cell persistence, and patients with durable response to therapy have relatively elevated levels of naïve and memory T cells (4,5,15-18). The concept of infusing T cells enriched in naïve and memory phenotypes is consistent with prior reports indicating naïve and/or memory T cells (T_N/MEM_ cells) enhance *in vivo* CAR-T cell function (19-21). Therefore, we hypothesized that engineering T_N/MEM_ cells to express our molecularly optimized CD19/CD20 bispecific CAR would yield a therapy, termed CART19/20, with strong efficacy coupled to improved CAR-T cell persistence and reduced CAR-T cell exhaustion. Here, we report the dose-escalation phase-1 trial evaluating CART19/20 in adults with relapsed/refractory NHL, and demonstrate durable responses and strong safety profile in patients treated with CART19/20 cells.

## RESULTS

### CAR construct and clinical trial design

We generated a CD19/CD20 bispecific CAR consisting of a single-chain variable fragment (scFv) derived from the anti-CD20 monoclonal antibody Leu16 fused to a second scFv derived from the anti-CD19 monoclonal antibody FMC63, followed by fusion of the scFv domains to the hinge domain of human IgG4, the transmembrane domain of human CD28, and the cytoplasmic signaling domains of human 4-1BB and CD3ζ (**Fig. 1A**) (6,7). The bispecific CAR was encoded by a third-generation self-inactivating lentiviral vector under the control of an elongation factor 1 alpha (EF1-α) promoter (22). We planned a phase-1 cell dose escalation trial with a fixed lymphodepletion chemotherapy of fludarabine 30 mg/m^2^ daily for three days and cyclophosphamide 500 mg/m^2^ daily for three days followed by CART19/20 infusion with dose levels of 50 × 10^6^ CAR^+^ T cells (DL1), 200 × 10^6^ CAR^+^ T cells (DL2), and 600 × 10^6^ CAR^+^ T cells (DL3), with each DL allowing ±30% range (**Fig. 1B**).

**Fig. 1.**
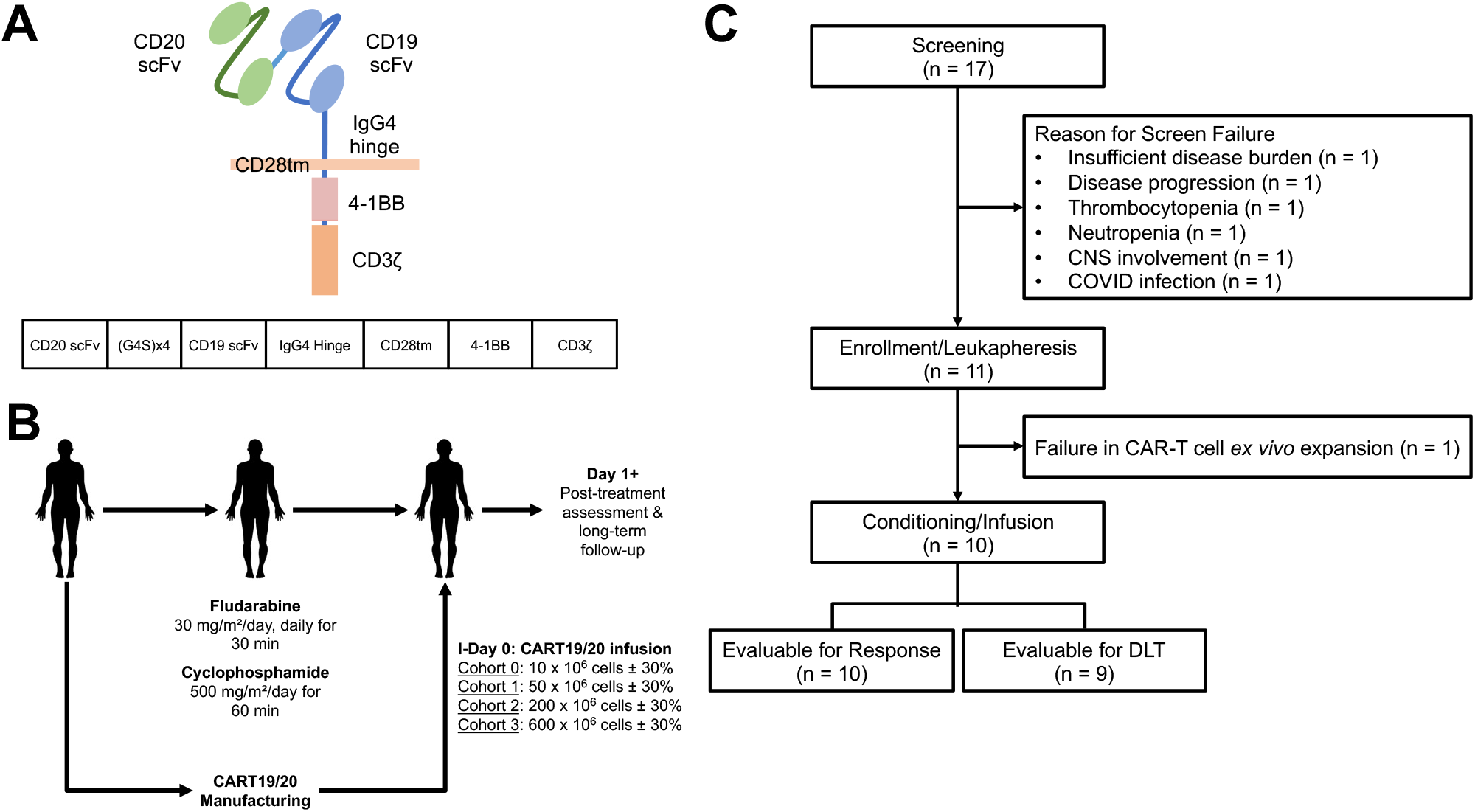
Design of phase-1 clinical trial evaluating CD19/CD20 bispecific CAR-T cell therapy (CART19/20) in patients with non-Hodgkin lymphoma, chronic lymphocytic leukemia, and small lymphocytic lymphoma. **(A)** Schematic of CD19/CD20 bispecific CAR construct. **(B)** Schematic of phase-1 dose escalation trial design. **(C)** Consolidated Standards of Reporting Trials (CONSORT) diagram of CART19/20 trial.

### Patient characteristics

Seventeen patients were screened, 11 patients went onto leukapheresis (**Fig. 1C**), and ten patients received CART19/20 infusion in two cohorts (DL1, n=7; DL2, n=3) (**Fig. 1C**). The median age at the time of CART19/20 infusion was 59 (range, 29–70) (**Table 1**). The diagnoses were mantle-cell lymphoma (MCL; n=1), follicular lymphoma (FL; n=3), *de novo* diffuse large B-cell lymphoma (DLBCL; n=1), transformed FL to DLBCL (n=3), primary mediastinal B-cell lymphoma (PMBCL; n=1), and high-grade B-cell lymphoma (HGBCL; n=1) with BCL6 and cMYC double-hit rearrangement. All patients with FL had progression of disease within 24 months after front-line treatment (POD24). The median lines of prior therapy were 3.5 (range, 2–4). One patient (Patient 004) was refractory to prior anti-CD19 bispecific T-cell engager (BiTE) therapy. All patients were CAR naïve and had stage-4 disease at the time of CART19/20 treatment. Nine patients were given bridging therapy prior to infusion due to progressive disease (**Table 1**).

**Table 1.** Patient demographics and treatment history.

| Patient ID | Age Range | Sex | Diagnosis | Prior Therapies |  |  | Disease Stage | ALC at Screening (x10<sup>3</sup>/μL) | Bridging Therapy | Dosing Level | CAR+ T Cells Received | CRS, (Grade) | ICANS (Grade) | Best Response (PFS) |
| --- | --- | --- | --- | --- | --- | --- | --- | --- | --- | --- | --- | --- | --- | --- |
|  |  |  |  | # of Lines | Therapies | Notes |  |  |  |  |  |  |  |  |
| 001 | 60–64 | M | MCL | 4 | •Bendamustine/rituximab<br>•Rituximab<br>•Acalabrutinib<br>•Umbralesib/ublituximab |  | IV | 0.40 | Cyclophosphamide | DL1 | 53 x 10<sup>6</sup> | Yes (1) | No | CR (30+) |
| 002 | 55–59 | M | FL | 4 | •Bendamustine/rituximab<br>•R-CHOP<br>•R-ICE<br>•Lenalidomide/rituximab | Primary Refractory | IV | 0.96 | •Cyclophosphamide<br>•R-EPOCH | DL1 | 58 x 10<sup>6</sup> | Yes (1) | No | CR (24+) |
| 003 | 25–29 | F | DLBCL (PMBCL) | 2 | •R-EPOCH<br>•R-ICE |  | IV | 0.17 | R-GEMOX | DL1 | 52 x 10<sup>6</sup> | Yes (1) | No | PD |
| 004 | 60–64 | M | FL | 4 | •Bendamustine/rituximab<br>•ROR1-Targeting Antibody-Drug Conjugate<br>•Anti-CD19 BITE<br>•Rituximab/gemcitabine/oxaliplatin | POD24; Refractory to CD19 BITE | IV | 1.02 | R-GEMOX (did not tolerate/complete rituximab) | DL1 | 56 x 10<sup>6</sup> | Yes (1) | No | CR (18) |
| 005 | 35–39 | F | FL | 3 | •Bendamustine/rituximab<br>•Rituximab<br>•Rituximab/dexamethasone | POD24 | IV | 1.12 | None | DL2 | 165 x 10<sup>6</sup> | Yes (1) | No | CR (18+) |
| 009 | 65–69 | M | DLBCL (NOS) | 4 | •Rituximab/cyclophosphamide/etoposide<br>•R-CHOP<br>•R-ICE<br>•ASCT | Primary Refractory | IV | 2.09 | GEMOX | DL2 | 158 x 10<sup>6</sup> | Yes (1) | No | CR (9+)* |
| 010 | 55–59 | M | DLBCL (tFL) | 3 | •Bendamustine/rituximab<br>•R-CHOP<br>•R-ICE |  | IV | 0.63 | ICE | DL1 | 42 x 10<sup>6</sup> | No | No | CR (12+) |
| 014 | 60–64 | M | DLBCL (tFL) | 2 | •Rituximab/gemcitabine/dexamethasone/carboplatin<br>•R-CHOP | Primary Refractory | IV | 2.60 | ICE | DL2 | 146 x 10<sup>6</sup> | No | No | CR (9+) |
| 016 | 70–74 | M | DLBCL (tFL) | 4 | •R-CHOP<br>•BR + polatuzumab<br>•R-ICE<br>•R-GEMOX |  | IV | 1.20 | GEMOX + R-GEMOX | DL1 | 37 x 10<sup>6</sup> | Yes (1) | No | PR (2) |
| 017 | 35–39 | M | DLBCL (HGBCL double-hit) | 2 | •R-CHOP<br>•R-EPOCH | Primary Refractory | IV | 1.88 | ICE | DL1 | 36 x 10<sup>6</sup> | No | No | PR (2+) |

\*Patient 009 elected to transition to comfort care shortly prior to the scheduled 12-month follow-up. Biopsy performed at that time showed no evidence of lymphoma. However, CR at 12 month was not verified with PET scan.

As of the data cutoff on July 11, 2022, a total of ten patients were evaluable for response. Nine patients were evaluable for dose-limiting toxicity (DLT), including six treated at DL1 and three treated at DL2. A decision was made to not escalate to DL3 based on the strong efficacy outcomes observed at the two lower dosing levels. The maximum tolerated dose was not reached.

### CART19/20 cell manufacturing and product characterization

Patient leukocytes harvested from leukapheresis were enriched for cells expressing CD62L, a marker for T_N/MEM_ cells, by magnetic bead-based cell separation. Leukapheresis products with greater than 5% CD14^+^ and/or CD25^+^ cells among viable singlets based on flow cytometry analysis were subjected to an additional CD14/CD25 depletion step to remove myeloid and regulatory T cells (Tregs), respectively, prior to CD62L enrichment. T_N/MEM_ cells were activated with an anti-CD3/CD28 colloidal nanomatrix-based activation reagent, lentivirally transduced to express the CD19/CD20 bispecific CAR, and expanded *ex vivo* for a total of 12 days (n=9),14 days (n=1, Patient 016), or 16 days (n=1, Patient 007) prior to cryopreservation to yield the CART19/20 product. Ten of the 11 products manufactured met release criteria, with one manufacturing failure due to low CAR^+^ T-cell counts that did not meet dose requirements (Patient 007). Patient 007 was diagnosed with stage-4 DLBCL transformed from lymphoplasmatic lymphoma. The patient had a low absolute lymphocyte count (ALC) of 0.16 × 10^3^ cells/μL at the time of screening, and the patient’s leukapheresis product was also low in white blood cell (WBC) count (32.79 × 10^3^/μL). Patient 007’s cells transduced and expanded poorly during *ex vivo* manufacturing (**Fig. 2A and B**), resulting in failure to meet the required CART19/20 cell dose for infusion.

**Fig. 2.**
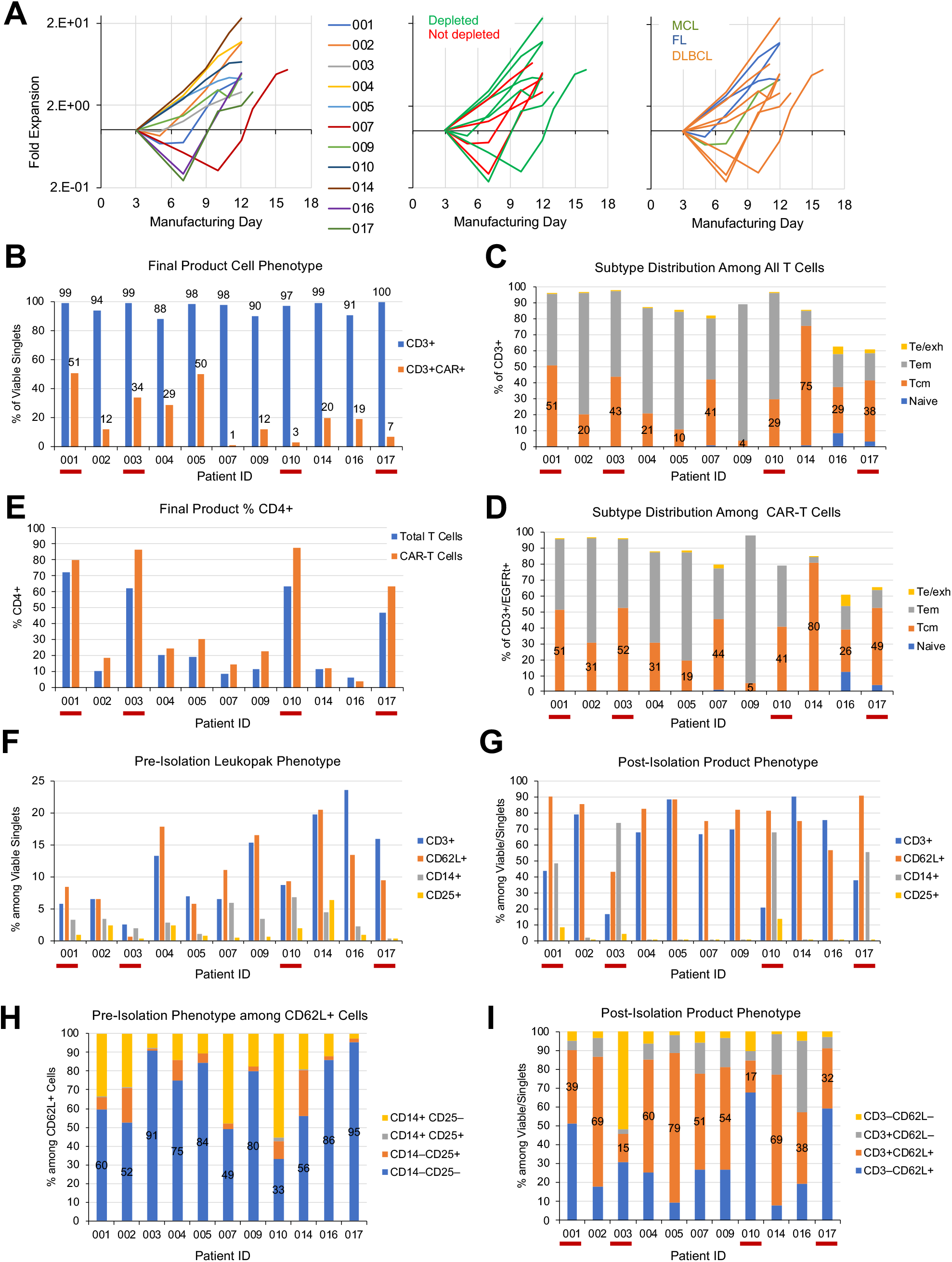
CART19/20 cells manufactured from naïve/memory T cells are enriched in memory phenotype. **(A)** Fold expansion of cell product during *ex vivo* manufacturing. Cell counts were normalized to counts on the day of transduction (day 3). Data are shown with color coding by patient (left), by whether starting cell population underwent CD14/CD25 depletion (middle), and by disease indication (right). **(B–D)** Flow cytometry performed on cryopreserved cell aliquots post thaw to characterize (B) CD3^+^ purity and transduction efficiency of final cell product, (C) T-cell subtype distribution among all CD3^+^ T cells, (D) T-cell subtype distribution among CAR-expressing T cells, and (E) % CD4^+^ among total T cells and CAR-expressing T cells. Te/exh: effector/exhausted T cells, CD45RA^+^/CD45RO^−^/CD62L^−^; Tem: effector-memory T cells, CD45RA^−^/CD45RO^+^/CD62L^−^; Tcm: central-memory T cells: CD45RA^−^/CD45RO^+^/CD62L^+^; naïve: CD45RA^+^/CD45RO^−^/CD62L^+^. **(F)** Phenotype of leukopak content prior to cell isolation. **(G)** Phenotype of cells obtained after isolation. **(H)** CD14 and CD25 expression patterns among CD62L+ cells in patient leukopak content prior to cell isolation. **(I)** CD3 and CD62L expression patterns among cells obtained after isolation. In panels B–G and I, red underscoring of patient ID indicates products that did not undergo CD14/CD25 depletion.

Overall, CART19/20 cell products contained a substantial fraction of central-memory T (Tcm) cells (median: 29.3%, range: 3.6%–74.9%; **Fig. 2C**), indicating the retention of memory phenotype in cell products manufactured from T_N/MEM_ cells. Of note, CAR-expressing T cells tend to have slightly higher Tcm content (median: 40.9%, range: 5.3%–80.1%) compared to the overall T-cell population (**Fig. 2D**). A breakdown of CD4^+^ vs. CD8^+^ subtype distribution reveals that CAR^+^ T cells tend to have higher % CD4^+^ than the total T-cell population (**Fig. 2E**), and CD4^+^ T cells tend to be more enriched in the Tcm phenotype compared to CD8^+^ T cells (**Supplementary Fig. S1A**).

#### Depletion of CD14/CD25 cells results in CD8-dominant T-cell products and no significant impact on ex vivo cell expansion, transduction efficiency, or memory phenotype distribution

We chose to incorporate CD14 depletion in order to minimize the presence of myeloid cells, which had been reported to reduce T-cell activation through phagocytosis of activation agents (23) and could potentially reduce transduction efficiency by competing with T cells for lentivirus uptake. The removal of immunosuppressive Tregs through CD25 depletion (24) aimed to further enhance the anti-tumor efficacy of CART19/20 products. The minimum threshold of ζ5% CD14^+^ and/or CD25^+^ cells for depletion was based on the empirical observation that up to 5% of antigen-positive cells can remain even after depletion during preclinical process development. Following this criterion, the leukapheresis products of Patients 001 and 003 were not subjected to depletion and proceeded directly to CD62L enrichment (**Fig. 2F**). However, the post-isolation cell population for Patients 001 and 003 showed a notable increase in % CD14^+^ (from 3% and 2% to 48% and 74%, respectively), together with an uptick in % CD25^+^ (from 0.9% and 0.3% to 8% and 4%, respectively; **Fig. 2G**). This unintended enrichment is consistent with the fact that a large fraction of CD62L^+^ cells in patient leukapheresis products are CD14^+^ and/or CD25^+^ (**Fig. 2H**), thus the CD62L enrichment step would simultaneously result in selective retention of CD14^+^ and/or CD25^+^ cells. Furthermore, the adherent nature of myeloid cells may also facilitate the retention of CD14^+^ cells during bead-based cell sorting in the absence of a depletion step. Based on these observations, the protocol was amended to trigger CD14/CD25 depletion when ≥5% of CD62L^+^ cells (as opposed to ≥5% of viable singlets) were CD14^+^ and/or CD25^+^, starting with the product for Patient 004.

Despite the fact that the post-isolation cell population for patients 001 and 003 contained a substantial number of myeloid cells, the CART19/20 cultures for these two patients showed typical fold expansion and viability levels during the manufacturing process (**Fig. 2A; Supplementary Fig. S1B**). Both final products exhibited high levels of CD3^+^ purity and CAR transduction efficiency (**Fig. 2B**), and had similar T-cell subtype distribution at the time of cryopreservation as products made from CD14/CD25-depeleted cells (**Fig. 2C and D**). The only clear difference was that products generated from non-depleted starting material were CD4 dominant, whereas products generated from CD14/CD25-depleted cells were CD8 dominant (**Fig. 2E**), potentially due to myeloid cells’ ability to stimulate CD4^+^ cells through MHC-II antigen presentation. Taken together, these results indicate the culture conditions succeeded in selectively expanding T cells while avoiding the negative impacts of myeloid cells observed in the manufacturing processes described in previous reports (23,25,26). Given that the starting % CD25^+^ was <1% in both Patients 001 and 003, there was minimal concern regarding Treg enrichment. Indeed, FOXP3 intracellular staining showed no clear correlation between CD25 depletion or lack thereof with the level of FOXP3 expression among CD4^+^ T cells in CART19/20 final products (**Supplementary Fig. 2**).

Among the subsequent products, two more were manufactured in the absence of CD14/CD25 depletion (Patients 010 and 017). Both products were enriched in CD4^+^ T cells, with no distinctive pattern in *ex vivo* expansion rate, transduction efficiency, or T-cell subtype distribution compared to CD14/CD25-depleted products (**Fig. 2B–E**), consistent with initial observations made with Patients 001 and 003.

#### Disease histology as risk factor in CAR-T cell manufacturing

Among the 11 manufacturing campaigns performed for this trial, three experienced a T-cell population contraction of >2 folds between the day of activation (day 1) and the day of transduction (day 3) (**Fig. 2A**). All three products were for patients diagnosed with DLBCL subtypes (Patients 007, 016, 017). Despite the reduced cell expansion, products for Patients 007, 016, and 017 had average T-cell subtype distribution (**Fig. 2C and D**). Transduction efficiency was low for two of the three products (**Fig. 2B**), and the product for Patient 007 failed to meet the dose-level requirement as previously noted. However, DL1 products were successfully manufactured for Patients 016 and 017 and administered to the patients.

### Safety and adverse events

Ten patients received CART19/20 cell products. One patient treated at DL1 (Patient 003) progressed prior to the end of DLT-monitoring period and was replaced per protocol and excluded from DLT evaluation. Infusions were generally well tolerated; 2 patients experienced a grade-1 infusion-related reaction (IRR) (**Table 2**). Grade-1 cytokine release syndrome (CRS) occurred in 6 patients; no grade 2 or higher CRS was observed in any patient. The median time from infusion to CRS was 8 days (range, 1-11) and the median duration was 2.5 days (range, 1-3). One dose of tocilizumab was given to Patient 009 for grade-1 CRS lasting greater than 48 hours. There were no cases of immune-effector cell associated neurotoxicity syndrome (ICANS), and no steroids were administered in the study for CRS or ICANS management. All patients experienced grade-3 or above adverse events consisting of generalized pain (n=2, 22%), hypotension (n=1, 11%), anemia (n=3, 33%), neutropenia (n=5, 56%), and thrombocytopenia (n=5, 56%). The only grade ≥3 adverse events attributable to CART19/20 cells were anemia, thrombocytopenia, and neutropenia experienced by Patient 009.

**Table 2.**
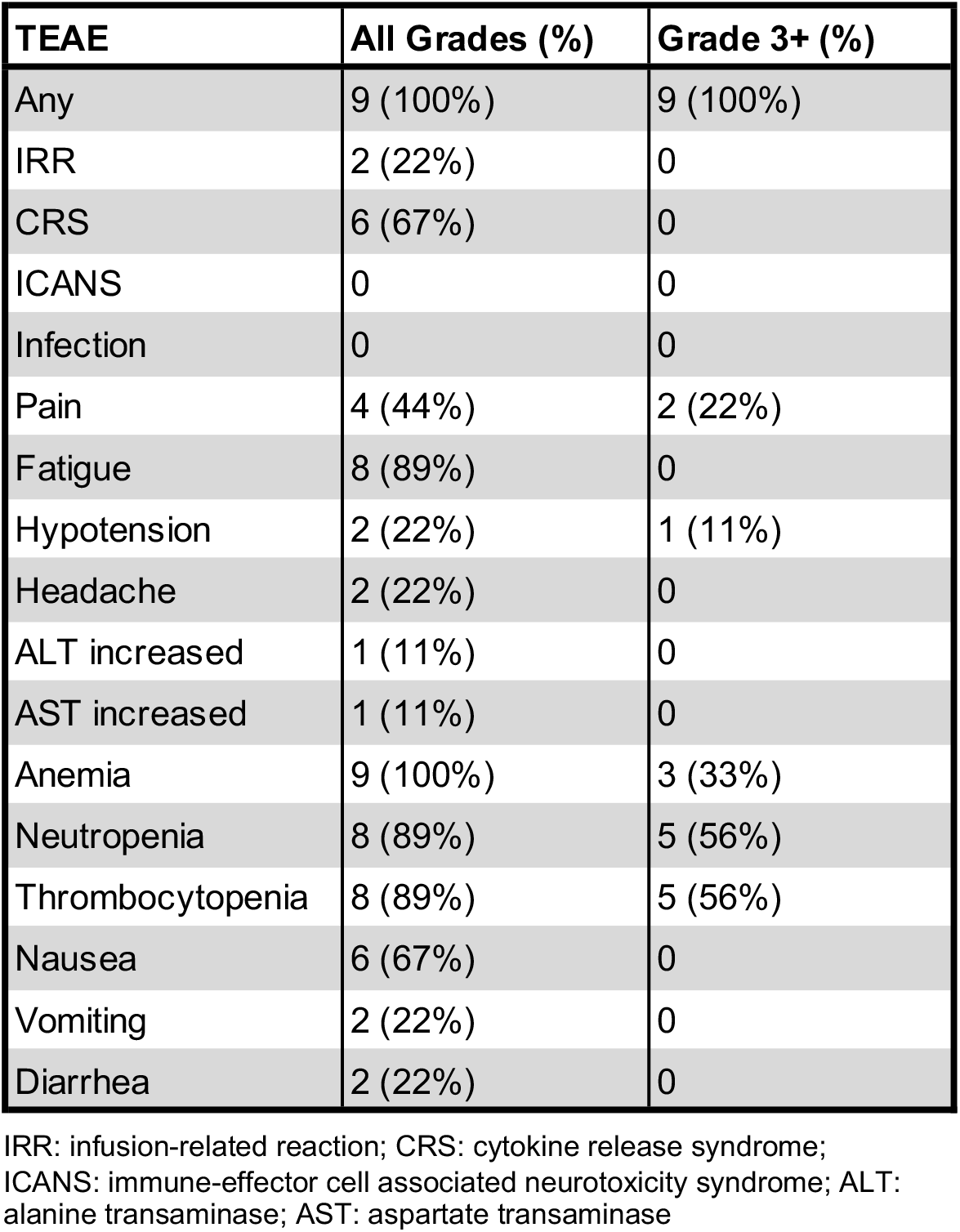
Adverse events.

In contrast to the other grade ≥3 hematologic adverse events, the cytopenia in Patient 009 persisted beyond the expected recovery time from lymphodepletion chemotherapy and resulted in the only DLT observed in this study. Patient 009 had received autologous stem-cell transplant (ASCT) 11 months prior to receiving CART19/20 cell infusion, and exhibited elevated levels of multiple cytokines—including IL-6, IFN-γ, IL-1RA, IL-1b, IL-2, IL-12, and GM-CSF— prior to CART19/20 cell infusion, suggesting baseline inflammation and potentially contributing to post-infusion cytopenia (**Supplementary Fig. S3A**). Post CART19/20 cell infusion, the patient experienced a typical, transient spike in C-reactive protein (CRP) and ferritin levels, but subsequently experienced a gradual increase in both that is unique among patients treated on this trial (**Supplementary Fig. S3B and C**). A bone marrow biopsy performed five months post CART19/20 cell infusion showed extensive, coalescing, non-necrotizing granulomatous inflammation. A trial of steroids was given with a transient improvement in pancytopenia. A repeat bone marrow biopsy performed ten months post CART19/20 cell infusion showed a hypocellular marrow with numerous histiocytes with increased hemophagocytic activity. Molecular characterization panel performed on this sample noted an expansion of the TET2 mutation to a variant allele frequency of 48% from a prior baseline of 1–3%, and a new ASXL1 mutation with a variant allele frequency of 65%. These findings suggest evolution of a myeloid neoplasm related to prior therapy.

Patient 009 elected to cease medical treatment shortly before the 12-month follow-up assessment. Given the severely hypocellular marrow and pancytopenia, T cells and peripheral mononuclear blood cells (PBMCs) could not be recovered in sufficient amounts to enable detailed follow-up analysis. Therefore, the patient’s grade 5 hypocellular marrow is considered possibly related to CAR-T cells and possibly related to preconditioning chemotherapy. Biopsy taken shortly prior to the patient’s election to end medical treatment showed no evidence of lymphoma.

No grade ≥3 adverse events attributable to CART19/20 cell therapy has been observed in any other patient treated on this trial.

### Response

Primary response assessment was performed 60 days after CART19/20 infusion by PET/CT scan. Nine out of ten patients responded to therapy (ORR = 90%). Seven out of ten patients achieved a CR by the first disease assessment at day 60 (CR rate = 70%), and two additional patients had a PR at day 60 (**Fig. 3 and 4A**). Bridging therapy did not result in significant reduction of tumor burden in the majority of cases (**Fig. 4A and Supplementary Fig. S4**), and there was no correlation between patient response and application of bridging therapy (**Table 1**). With a median follow-up of 17 months from time of CART19/20 cell infusion (range 2– 30 months), the median overall survival (OS) and progression-free survival (PFS) were not reached (**Fig. 4B**).

**Fig. 3.**
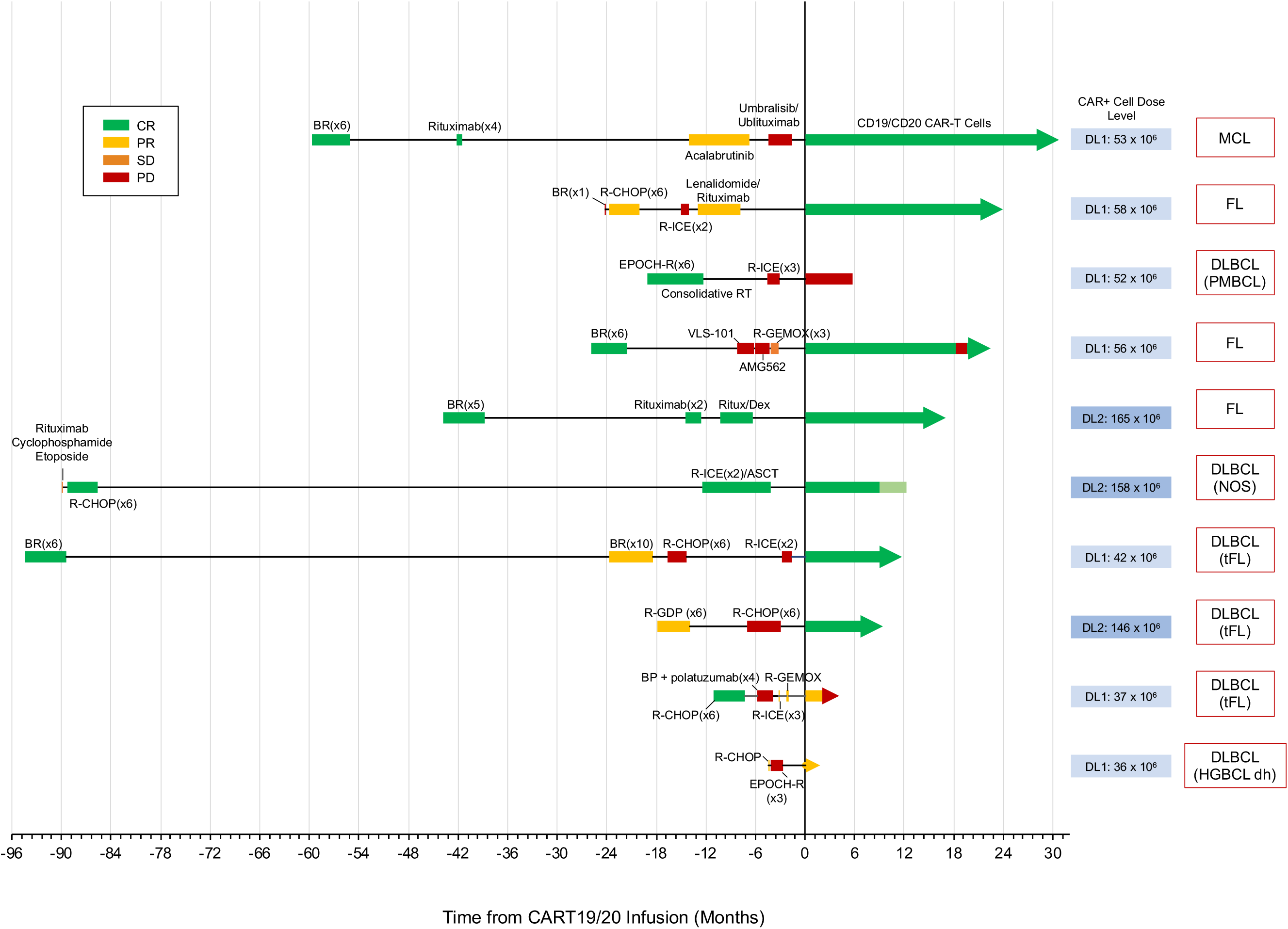
Patients refractory to multiple prior lines of treatment respond to CART19/20 cell therapy. Timeline of individual patients’ response to prior treatment and to CART19/20 cell therapy. The disease indication and dose of CART19/20 cells received are also indicated for each patient. MCL, mantle-cell lymphoma; FL, follicular lymphoma; PMBCL, primary mediastinal B-cell lymphoma; DLBCL, diffuse large B-cell lymphoma; tFL, transformed follicular lymphoma.

**Fig. 4.**
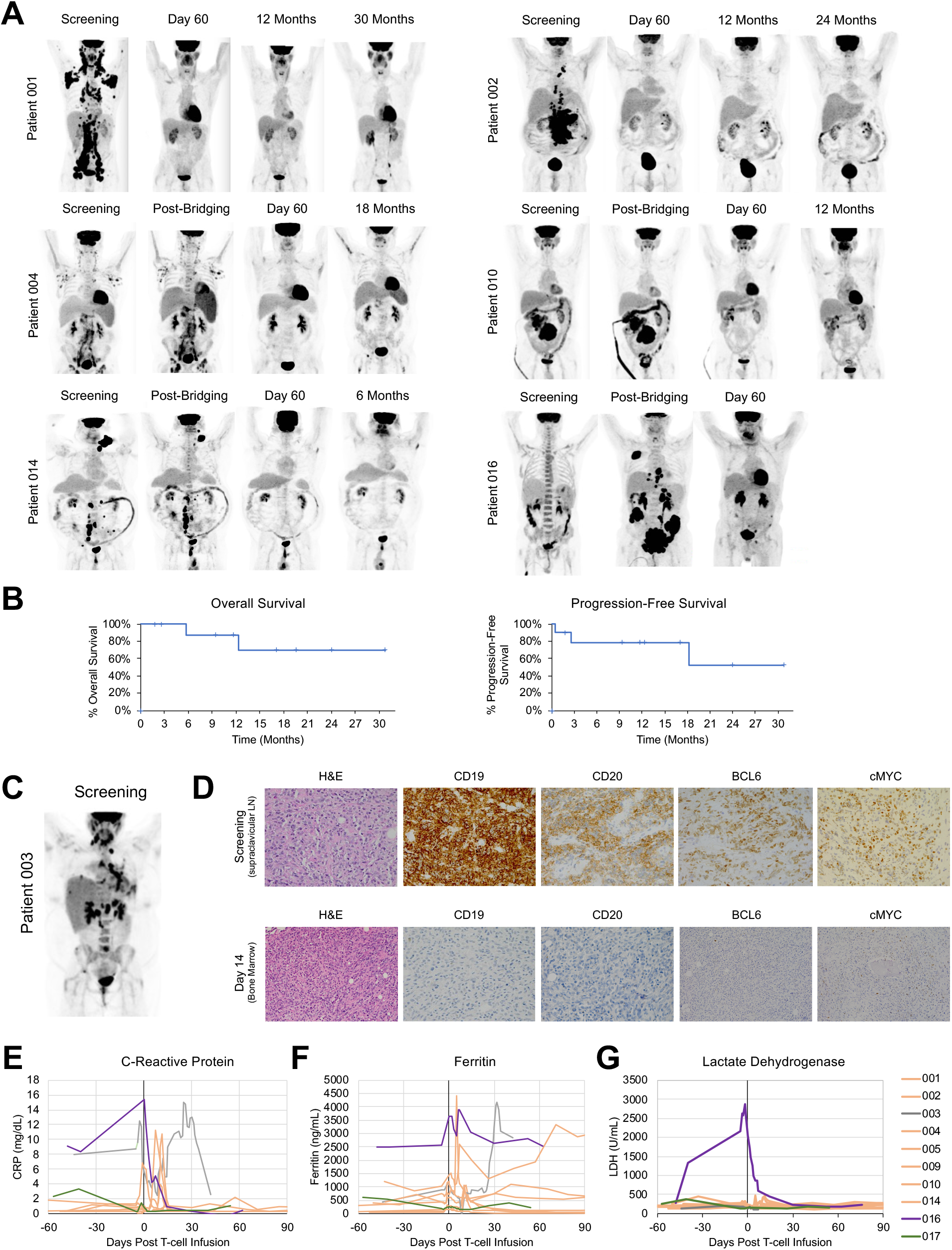
CART19/20 cell therapy is highly effective in treating relapsed/refractory non-Hodgkin lymphomas. **(A)** Representative PET scans of patients treated with CART19/20. **(B)** Overall survival and progression-free survival curves from the time of CART19/20 cell infusion. **(C)** PET scan obtained at screening for Patient 003, indicating pulmonary involvement of PMBCL. **(D)** Immunohistochemistry (IHC) analysis of Patient 003 tissue biopsies; original magnification x160. Supraclavicular lymph node biopsy obtained at screening and bone-marrow biopsy obtained 14 days post CART19/20 infusion were analyzed by IHC. Results reveal rapid emergence of a CD19^−^CD20^−^BCL6^−^cMYC^−^ tumor population within 14 days of CART19/20 treatment. **(E–G)** C-reactive protein (CRP), ferritin, and lactate dehydrogenase (LDH) levels of all patients treated with CART19/20 cell therapy.

The one patient who did not respond to therapy (Patient 003) was diagnosed with PMBCL (**Fig. 4C**), and had low ALC at the time of screening (0.17 × 10^3^ cells/μL). The patient exhibited marked clinical improvements, including reduced dyspnea and pain, in the week after CART19/20 cell infusion and was discharged from the hospital. However, disease progression was detected 14 days post CART19/20 infusion in a bone-marrow biopsy performed as part of routine evaluation for CAR-T cell infiltration and expansion. The screening biopsy of a supraclavicular lymph node from Patient 003 was positive for CD19, CD20, CD30 (patchy), BCL2, BCL6, and cMYC (**Fig. 4D**), with kappa light chain restriction; the screening biopsy was negative for CD10 and BCL1. In contrast, the bone-marrow sample obtained 14 days after CART19/20 infusion expressed CD30 and weak BCL2, and was negative for CD10, CD19, CD20, BCL1, BCL6 and cMYC (**Fig. 4D**), indicating a clonal shift in the tumor population.

To further understand the mechanism of dual antigen loss after CART19/20 treatment, an additional biopsy of the lung was obtained and analyzed by bulk RNA sequencing (RNA-seq). Consistent with the immunohistochemistry and flow cytometry results, RNA-seq showed loss of gene expression for CD19, CD20 and BCL6 (reads per kilobase transcript, per million mapped reads (RPKM) = 0.12, 0.11, and 0.72, respectively), and low cMYC expression (RPKM = 2.04). RNA-seq data further revealed a lack of CD22 expression (RPKM = 0.07). Transcriptomic data interpretation is limited due to the lack of pre-treatment tissues for RNA-seq analysis. Nevertheless, the concomitant loss of BCL6 and cMYC—two antigens not under selective pressure from the CD19/20 bispecific CAR—within 14 days of CART19/20 cell infusion suggests the possibility of heterogenous tumor that contained a pre-existing antigen-negative subclone.

Patient 016, who was diagnosed with tFL, achieved a PR at day 60 (**Fig. 4A**) but progressed on day 90. Both Patients 003 and 016 exhibited elevated CRP prior to CART19/20 cell infusion (**Fig. 4E**), and both had elevated ferritin levels as of the last assessed time point (**Fig. 4F**). Following bridging therapy and prior to CART19/20 cell infusion, Patient 016 developed extremely high levels of lactate dehydrogenase (LDH) (**Fig. 4G**). The low ALC observed in Patient 003 and high pre-leukapheresis LDH observed in Patient 016 have both been shown to correlate with low odds for CR after CAR-T cell therapy (27). In contrast, Patient 017, who similarly achieved a PR at day 60, exhibited normal patterns of CRP, ferritin, and LDH levels (**Fig. 4E–G**), as well as a pre-leukapheresis ALC within normal range (**Table 1**). This patient was diagnosed with primary-refractory HGBCL with BCL6 and cMYC rearrangement, and remains in PR as of data cutoff.

The seven patients who achieved a CR include one patient diagnosed with MCL, three patients with DLBCL (one *de novo* and two tFL) and three patients with FL (**Fig. 3**). All patients with FL were POD24, and the majority of patients were characterized by high tumor burdens (**Supplementary Fig. S4**). Among the seven patients who achieved a CR, three (Patients 002, 009, and 014) had primary refractory disease, and a fourth (Patient 004) was refractory to anti-CD19 BiTE therapy (**Table 1**). In addition to anti-CD19 BiTE, Patient 004 was also refractory to ROR1-targeted antibody-drug conjugate therapy and progressed through the 2^nd^–4^th^ lines of therapy within 5 months (**Fig. 3**). Flow cytometry analysis of Patient 004’s peripheral blood at the time of screening indicated the presence of CD20^+^CD19^dim/–^ cells (**Fig. 5A**). This population was substantially reduced within 7 days of CART19/20 infusion, confirming CART19/20’s ability to target tumor cells that have downregulated or lost CD19 expression. Patient 004 achieved a CR within 60 days and remained in CR until month 18, when reemergence of CD20^+^CD19^+^ FL was detected. Given the patient’s history of therapy resistance and 18-month response to CART19/20, permission was obtained from the FDA to re-dose Patient 004 with 126 × 10^6^ CART19/20 cells, using extra aliquots of the same cell product as the first infusion. At day 60 post re-dosing, the patient again achieved a CR and remains in CR as of data cutoff (**Fig. 5B**).

**Fig. 5.**
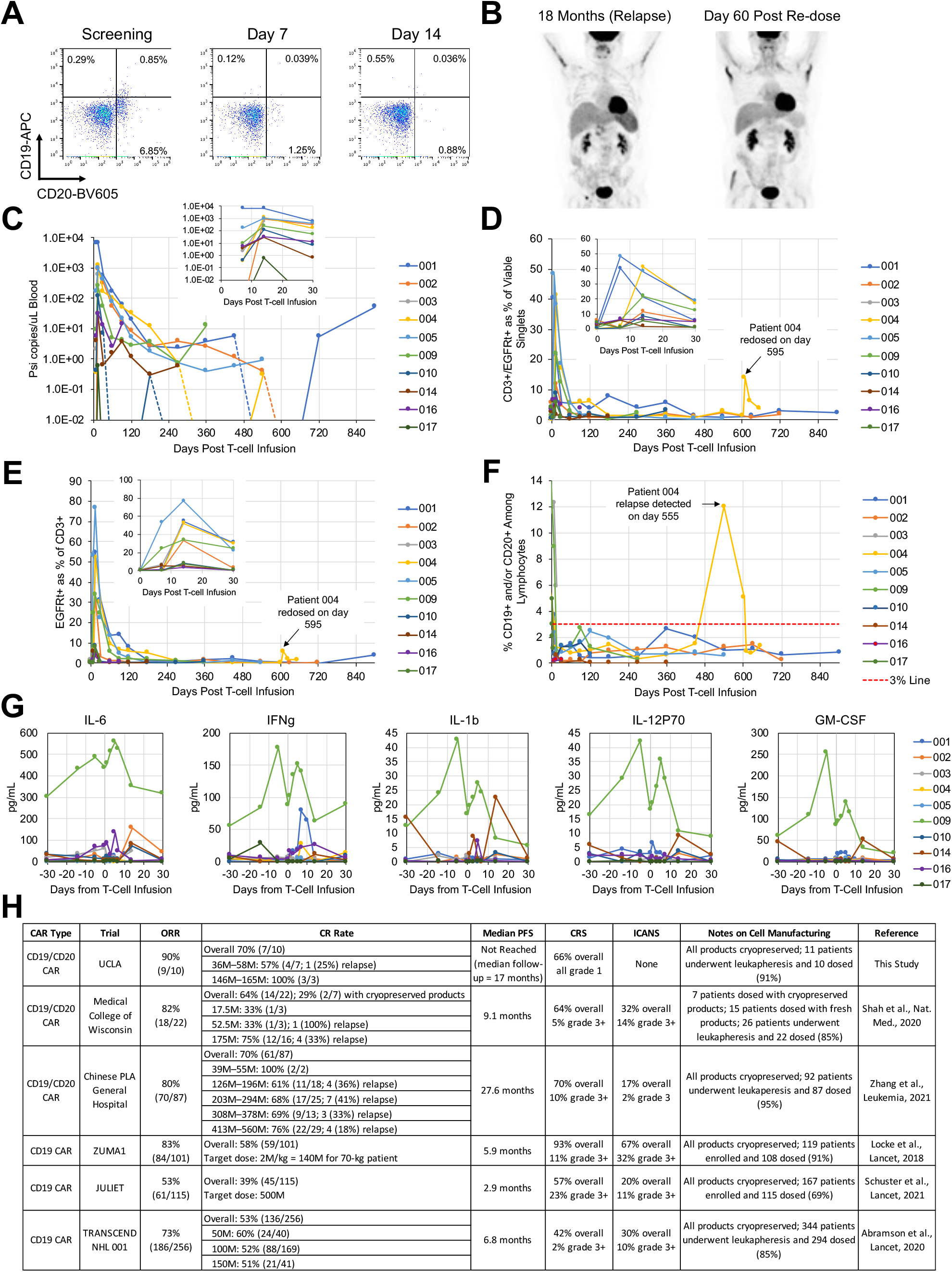
CART19/20 cells exhibit sustained persistence and efficacy with strong safety. **(A)** Flow cytometry analysis on peripheral blood samples collected from Patient 004 at screening as well as 7- and 14-days post CART19/20 infusion. CD19 and CD20 surface staining results indicate the presence of CD20^+^CD19^dim/–^ cells in Patient 004 prior to CART19/20 cell treatment. PET scans of Patient 004 at the time of relapse and at 60 days post second dose of CART19/20 cells. **(C)** Presence of CAR transgene as quantified by droplet digital PCR. The psi signal integrated through lentiviral transduction was quantified. Inset shows zoomed-in data from the first 30 days post CART19/20 infusion. **(D)** Presence of CAR-expressing T cells among peripheral blood mononuclear cells (PBMCs) as quantified by flow cytometry. **(E)** Presence of CAR^+^ cells among T cells in patient peripheral blood as quantified by flow cytometry. **(F)** Presence of CD19^+^ and/or CD20^+^ cells among lymphocytes as quantified by flow cytometry. A 3% line is shown to denote threshold for B-cell aplasia. **(G)** Serum cytokine levels as qualified by Luminex multiplex assay. **(H)** Comparison of CART19/20 cell therapy results with reported data from trials evaluating other CD19/CD20 bispecific CAR-T cell therapies and commercially available CD19 CAR-T cell therapies.

### CART19/20 cell persistence and B-cell aplasia

The presence of CART19/20 cells in peripheral blood post infusion were detected by both flow cytometry and droplet digital PCR (ddPCR). CAR copy number quantified by ddPCR consistently peaked at 14 days post infusion (**Fig. 5C**). At peak expansion, up to 48% of all viable singlets in PBMCs were CAR^+^ T cells (median 11.6%; range 1.5%–48.3%; **Fig. 5D**), and up to 77% of all CD3^+^ T cells were CAR^+^ (median 33.8%; range 3.7%–76.6%; **Fig. 5E**). At the time of data cutoff, all seven patients who were in PR (n=1) or CR (n=6) remained in B-cell aplasia based on flow cytometry analysis of peripheral blood (**Fig. 5F**), indicating functional persistence of CART19/20 cells.

For Patient 004, who remained in CR for 18 months before relapsing, CAR^+^ T cells dropped below ddPCR detection at 12 months but reemerged at the time of relapse (**Fig. 5C**), suggesting the presence of a residual CART19/20 cell population that expanded upon tumor relapse but was insufficient to prevent tumor outgrowth. The patient experienced a second wave of CAR^+^ T-cell expansion after receiving the second dose of CART19/20 cells (**Fig. 5D,E**). The more modest expansion post re-dosing compared to the initial peak suggests the possibility of resistance development toward the CAR-T cell product, but the re-dose was nonetheless successful in re-establishing a CR for this patient. The lower tumor burden and thus lower antigen load at relapse compared to initial screening (**Figs. 4A** and **5B**) could also have contributed to the lower CAR-T cell expansion upon re-dose. B cells were undetectable in the peripheral blood of Patient 004 after the first CART19/20 cell infusion, was detected at the time of relapse, and dropped below detection again after receiving the second dose of CART19/20 cells, consistent with response to therapy (**Fig. 4F**).

As previously noted, CART19/20 cell products were either CD4- or CD8-dominant at the time of cryopreservation, depending on whether CD14/CD25 depletion was performed (**Fig. 2E**). Interestingly, regardless of the CD4:CD8 ratio in the cryopreserved cell product, Both CAR-expressing T cells and total T cells rapidly became CD8-dominant after CART19/20 cell infusion in all but one patient (**Supplementary Fig. S5**). The CD8 dominance declines over time, mirroring recently reported findings in a decade-long follow up of two patients with leukemia treated with CD19 CAR-T cell therapy (28).

### Cytokine assessments

Consistent with clinical presentation of grade 1 CRS for over 48 hours, Patient 009 showed elevated levels of most cytokines in peripheral blood compared to other patients in the trial (**Fig. 5G; Supplementary Fig. S3A**). As a whole, cytokine levels observed in patients treated with CART19/20 cells are similar to or substantially lower than values reported in earlier trials for single-input CD19 CAR-T cell therapies (3,5,29-31) and CD19/CD20 bispecific CAR-T cell therapies (18,32). The relatively low peak cytokine levels in patients treated with CART19/20 may be a contributing factor to the strong safety profile observed in this trial to date. Taken together, results of this phase-1 clinical trial indicate CART19/20 cell therapy is safe and effective for the treatment of NHL (**Fig. 5H**).

## DISCUSSION

CD19 CAR-T cell therapy became the first FDA-approved gene-modified cell therapy in 2017, and is making rapid progress toward incorporation in earlier lines of treatment for B-cell malignancies. However, antigen escape and lack of T-cell expansion and persistence remain key factors that limit the frequency as well as durability of response in patients treated with CD19 CAR-T cell therapy (1-5,15-17,33,34). Early results of our phase-1 trial on CART19/20 cell therapy demonstrate that dual-targeting of CD19 and CD20 using a molecularly optimized single-chain bispecific CAR (6,7) is safe and effective in patients with relapsed/refractory NHL, with potentially higher CR rates and lower toxicity using lower T-cell doses compared to previously reported clinical candidates (18,35).

In this trial, ten patients were treated with a median dose of 55 × 10^6^ CART19/20 cells, with no patient receiving more than 165 × 10^6^ cells. These dosing levels are substantially lower than the median dosage level used in clinical trials evaluating other CD19/CD20 bispecific CAR clinical candidates (18,35) as well as in pivotal trials for single-input CD19 CAR-T cell products including axicabtagene ciloleucel (36,37), tisagenlecleucel (38,39), and lisocabtagene maraleucel (40). Despite the low dosage used, strong efficacy was observed in this trial (90% ORR; 70% CR rate), with a heavily pretreated patient population carrying high tumor burden and highly aggressive disease. To date, all but one patient who achieved a CR has remained in CR. The only patient to experience a relapse did so after 18 months, and the patient subsequently returned to CR after receiving a second dose of CART19/20 cells. With a median follow-up of 17 months, median PFS has not been reached in this trial.

Importantly, the strong efficacy observed in this trial was achieved with no ICANS of any grade and no CRS above grade 1 after CART19/20 infusion. To date, the vast majority of grade ≥3 adverse events experienced by patients treated with CART19/20 were toxicities attributable to bridging and lymphodepletion chemotherapies in patients with substantial previous chemotherapy exposure. One patient death (Patient 009) from grade 5 hypocellular marrow is considered possibly related to treatment, and possibly related to myeloid neoplasm consequent to therapies received prior to CART19/20. Although no evidence of lymphoma was detected in the patient at the time of elected transition to comfort care, this case underscores the potential benefit of providing CAR-T cell therapy as an earlier line of treatment, which could reduce the total amount of chemotherapy exposure and related toxicities to the patient. Overall, the safety profile observed in this trial compares favorably with prior reports of CD19-targeted as well as CD19/CD20-targeted CAR-T cell therapies (18,35-41), and supports the possibility of combining strong efficiency with a high level of safety.

The trial reported here included one patient with MCL, three patients with FL, and five patients with DLBCL of various subtypes. It is plausible that different DLBCL subtypes have different response rates to CAR-T cell therapy, although no trial reported to date has been statistically powered to evaluate the responses of different DLBCL subtypes to CD19 CAR-T cell therapy (36,39,40). FL is often described as an indolent disease, and has shown favorable response rates to therapy (38). It should be noted that in this trial, all three patients with FL were POD24, including one patient who was primary refractory (Patient 002), and another patient who was refractory to the three lines of therapy immediately before CART19/20, including CD19-targeted BiTE (Patient 004). Therefore, the subjects enrolled in this trial represented a particularly challenging subset of patients with FL, and all achieved a CR post CART19/20 cell infusion. In addition to four other patients with MCL or DLBCL who also achieved a CR, one patient with primary refractory, double-hit HGBCL (Patient 017) achieved a PR at day 60. This patient will continue to be monitored for the possibility of a delayed CR. Overall, CART19/20 cell therapy has shown robust efficacy in a highly pretreated patient population with challenging disease profiles.

The only patient who did not respond to CART19/20 therapy to date (Patient 003) had PMBCL refractory to R-ICE salvage chemotherapy. Despite notable clinical improvement in the week post CART19/20 infusion, this patient experienced a rapid emergence of CD19^−^CD20^−^ tumor cells that had also lost BCL6 and cMYC expression within 14 days of CART19/20 treatment. The number of protein-expression changes combined with the rapidity of clonal shift suggests a pre-existing tumor subpopulation that was able to swiftly expand after CART19/20 cells eliminated the originally dominant CD19^+^CD20^+^ tumor cells. The emergence of CD19^−^ tumor was previously reported in the relapse of a patient with PMBCL treated with CD19 CAR-T cell therapy, and sequencing analysis indicated the likely cause was a pre-existing clone that expanded under CD19-targeted selective pressure (42).

Promoting T-cell persistence and function through the generation of cell products enriched in naïve and memory cell types was a key element of the CART19/20 therapy design. CART19/20 products were generated from a T_N/MEM_ cell population obtained through bead-based enrichment of CD62L expression, and the final products retained substantial central-memory T-cell content. Contrary to our original expectations, the presence of the CD14^+^ cells did not adversely impact our ability to successfully manufacture CART19/20 cell products with high T-cell purity and clinical efficacy. The only clearly measurable impact of CD14^+^ cell presence was the CD4:CD8 ratio of the final CART19/20 product, with the presence of CD14^+^ cells leading to CD4-dominant products while the depletion of CD14^+^ cells from the starting material led to CD8-dominant products. However, based on the results to date, there is no correlation between the CD4:CD8 ratio and clinical outcome. Similarly, the lack of Treg depletion through CD25 did not show measurable impact on treatment outcome, which is consistent with prior reports (43).

A key question of interest is how the CART19/20 cells evaluated in this trial are able to achieve the high level of efficacy at low dosage level and without incurring the type of toxicities observed in comparable trials. As previously reported, the CD19/CD20 bispecific CAR used here had been optimized at a sequence level to maximize efficacy (6). Safety was not a consideration in the CAR engineering process. However, the robust efficacy enabled the use of a very low cell dose to achieve therapeutic benefit, and the low cell dose may in turn have contributed to the favorable safety profile observed in this trial. Unlike conventional CD19 CAR-T cell therapy, CART19/20 cells are capable of eliminating CD19^dim/–^ tumor cells within 7–14 days of T-cell infusion (**Fig. 5A**). In addition to safeguarding against antigen escape, the rapidity with which CART19/20 eliminates tumor cells may contribute to the limited toxicity observed— i.e., the bulk of the tumor could be eliminated before T cells reach peak expansion at 14 days post infusion, thus providing temporal separation between high tumor burden and large CAR-T cell numbers in the patient. Finally, the use of T_N/MEM_-derived cells may further contribute to the potency and safety profile of CART19/20 cells by reducing peak cytokine levels while retaining long-term anti-tumor efficacy.

In summary, early results from this phase-1 trial indicate that autologous, CD19/CD20 dual-targeting CAR-T cells enriched in naïve and memory phenotypes are safe and highly efficacious in the treatment of relapsed/refractory NHL. Our results suggest potent clinical efficacy can be achieved while avoiding severe toxicities typically associated with CAR-T cell therapy, and highlight the utility of dual-antigen targeting cell-based immunotherapy.

## METHODS

### Trial Design

A prospective, first-in-human phase I clinical trial assessing CART19/20 in adult patients with relapsed/refractory NHL and CLL/SLL was initiated at the University of California, Los Angeles (UCLA) Medical Center. The study was approved by the UCLA Institutional Review Board (IRB) and registered with ClinicalTrials.gov (NCT04007029). Informed written consent was obtained in accordance with the Declaration of Helsinki, International Conference on Harmonization (ICH) Good Clinical Practice (GCP), US Code of Federal Regulations for Protection of Human Subjects, the Health Insurance Portability and Accountability Act, and local regulations. Data monitoring was conducted by the UCLA Jonsson Comprehensive Cancer Center (JCCC) Data Safety and Monitoring Board (DSMB). The primary endpoint was safety, defined by the incidence and severity of dose-limiting toxicities (DLTs), as well as determination of the maximum tolerated dose (MTD). Secondary endpoints included clinical efficacy measures, and analysis of CAR T-cell persistence and B-cell aplasia. Exploratory endpoint was cytokine release syndrome analysis.

### Patient enrollment and eligibility

Patients eligible for the clinical trial were ≥18 years old with diffuse large B-cell lymphoma (DLBCL) or primary mediastinal B-cell lymphoma (PMBCL) after ≥2 prior lines of therapy, or with mantle-cell lymphoma (MCL), follicular lymphoma (FL), chronic lymphocytic leukemia (CLL) or small lymphocytic lymphoma (SLL) after ≥3 prior lines of therapy. Transformed indolent lymphomas, including Richter transformation, were eligible and previous lines of therapy were considered from the time of transformation. Autologous stem cell transplant (ASCT) recipients were allowed in the study. Patients were required to have greater than 30% positivity in malignant cells of CD19 and/or CD20, as well as measurable tumor burden on PET/CT. Any NHL- or CLL/SLL-directed therapy, including corticosteroids, within 14 days of initiation of lymphodepletion chemotherapy was exclusionary. After leukapheresis, bridging therapy was permitted at the investigator’s discretion. Lymphodepletion chemotherapy, consisting of fludarabine 30 mg/m^2^ and cyclophosphamide 500 mg/m^2^, was administered on day −5 through day −3 before infusion.

### Toxicity assessment

Adverse events were recorded for all treated patients until disease relapse or death, with incidence and severity graded using the Common Terminology Criteria for Adverse Events (CTCAE) version 5.0. CRS was graded according to the ASTCT and Lee criteria, with the former guiding treatment (44), For neurotoxic events, the ASTCT criteria was for scoring and treatment, with specific guidance to key disorders outlined by Neelapu et al. (45).

### Response assessment

The clinical response in lymphoma was evaluated with the criteria defined by The Revised Cheson Response Criteria and Lugano Classification (46,47). The overall response rate (ORR) was defined as the total of complete responses (CR) and partial responses (PR).

### CART19/20 cell manufacturing

Fresh patient leukapheresis products were analyzed by flow cytometry to determine the CD3^+^, CD62L^+^, CD14^+^, and CD25^+^ cell frequency. When needed, cells were labeled with anti-CD14 and anti-CD25 CliniMACS microbeads and depleted using the CliniMACS Plus system (Miltenyi Biotec). Remaining cells were subsequently enriched for CD62L using the same system to yield T_N/MEM_ cells. T_N/MEM_ cells were activated with TransAct (Miltenyi Biotec) and transduced with GMP-grade lentivirus. Patient cells were expanded *ex vivo* for a total of 12–16 days prior to cryopreservation.

### Flow cytometry analysis of lymphocytes

For pre- and post-isolation leukapheresis product analysis, samples were stained with antibodies for CD3, CD14, CD25, and CD62L. For CART19/20 final product analysis, cryopreserved products were thawed, washed with PBS, and stained with antibodies for CD3 and epidermal growth factor receptor (EGFR). The CD19/CD20 bispecific CAR is co-expressed with a truncated, non-signaling EGFR (EGFRt), thus EGFRt serves as a proxy for CAR expression. For patient peripheral blood analysis, blood samples were collected in ethylenediaminetetraacetic acid (EDTA) tubes, and peripheral blood mononuclear cells (PBMCs) were collected using the SepMate system (STEMCELL Technologies) following manufacturer’s protocol. Isolated PBMCs were frozen until use. Thawed cells were surface stained with antibody panels for T-cell phenotype (CD3, CD4, CD8, CD62L, CD45RA, CD45RO, and EGFR), or B-cell quantification (CD19, CD20, CD56, CD3, CD14, and SYTOX Blue). Flow cytometry was performed on an Attune NxT flow cytometer (ThermoFisher), and data were analyzed using FlowJo v.10.7.1 (FlowJo, LLC). Gating strategies are shown in Extended Data Figure 4.

### Cytokine analysis

Patient peripheral blood was collected into red-top tubes containing no anti-coagulant or preservative, allowed to clot for 30 minutes in the upright position at room temperature, transferred to a conical tube, and centrifuged at 900 x g for 10 minutes. The supernatant was frozen in aliquots until use. Cytokine analysis was performed by the UCLA Immune Assessment Core Facility using the Luminex 38-plex human cytokine chemokine panel, following manufacturer’s protocols.

### Statistical analysis

Descriptive statistics were measured by median and range for continuous variables and counts and percentages for categorical variables. Patients were censored at the time of the last follow up. Duration of remission (DOR), progression-free survival (PFS), and overall survival (OS) were estimated by Kaplan-Meier method. The statistical software packages used was IBM SPSS statistics. DOR was defined as the time of the first documented CR/PR until the first date that recurrent or progressive disease is objectively documented, or until death. PFS was defined as the time of CART19/20 infusion until documentation of objective disease progression or death due to any cause. OS was measured from the date of CART19/20 infusion in the clinical trial until death.

## Data Availability

All data produced in the present study that are legally eligible for release are available upon reasonable request to the authors.

## ACKNOWLEDGEMENTS

We thank the study participants and all members of the Cellular Therapy Program at UCLA. This work was supported by the Parker Institute for Cancer Immunotherapy (grant no. 20163828 to Y.Y.C) and Jean and Stephen Kaplan (gift to Y.Y.C.). This trial was additionally supported by the Aramont Clinical/Translational Research Program in Hematologic Malignancies and the Hornsey Foundation. A.R. is supported by NIH grants R35CA197633 and P30CA016042. J.M.T. is supported by the Jaime Erin Follicular Lymphoma Research Consortium. This study utilized the UCLA Jonsson Comprehensive Cancer Center (JCCC) Flow Cytometry Shared Resource and Technology Center for Genomics & Bioinformatics, which are supported by the NIH Cancer Center Support Grant (grant no. P30CA016042 to Michael A. Teitell). We thank the UCLA Human Gene and Cell Therapy Facility (HGCTF) for performing Quality Assurance review of our Good Manufacturing Practice (GMP) cell-manufacturing process. We thank Dr. Bea Fernandez her assistance in performing vector copy number (VCN) and lentivirus titer analyses. We thank Dr. Xiangzhi Meng and Dr. Ximin Chen for assistance in performing bulk RNA-seq data analysis. We thank the UCLA Immune Assessment Core for performing cytokine analysis.

## AUTHOR CONTRIBUTIONS

S.M.L., Y.Y.C., and A.R. conceived and designed the study. Y.Y.C., S.M.L., C.M.W., B.J., S.N.G., J.N., J.T., J.M.C., M.R., B.B.M., C.H., M.K.G., T.S, S.B.G., M.A., J.S., A.A., K.N., M.M., S.D.V., P.Y., C.O., G.J.S., and J.M.T. collected and assembled the data. Y.Y.C., S.M.L., M.A., J.S., B.J., C.H., M.K.G., T.S, A.A., and A.R. analyzed and interpreted the data. Y.Y.C., S.M.L., S.N.G., and A.R. wrote the manuscript.

