## Supplemental Figures 1-5 for "CD19/CD20 Bispecific Chimeric Antigen Receptor (CAR) in Naïve/Memory T Cells for the Treatment of Relapsed or Refractory Non-Hodgkin Lymphoma"

Sarah M. Larson, et al.

**This file includes:**

Figs. S1 to S5
Captions for Figs. S1 to S5

### Supplementary Figure S1

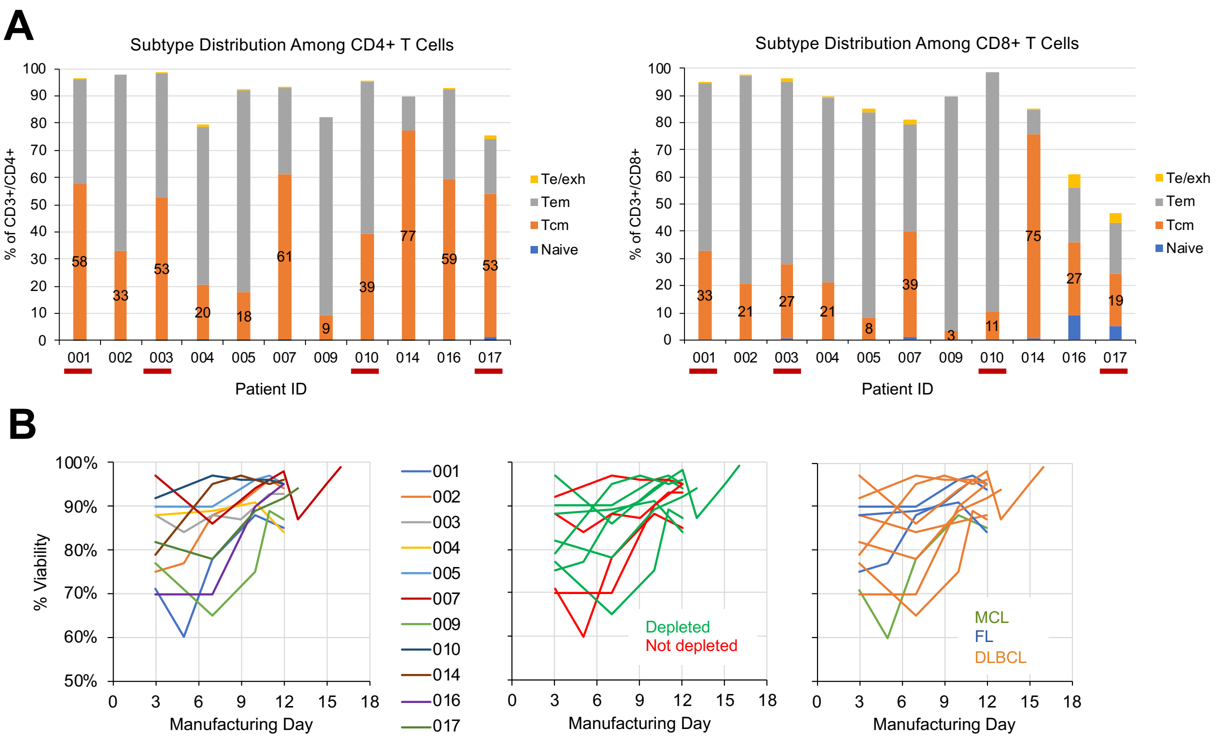


**Supplementary Fig. S1. CART19/20 cell product characteristics. (A)** T-cell subtype distribution in cryopreserved CART19/20 cell products, with data segregated for CD4$^{+}$ and CD8$^{+}$ populations. Results indicate CD4$^{+}$ T cells are more enriched in the central-memory phenotype compared to CD8$^{+}$ T cells. Te/exh: effector/exhausted T cells, CD45RA$^{+}$/CD45RO$^{-}$/CD62L$^{-}$; Tem: effector-memory T cells, CD45RA$^{-}$/CD45RO$^{+}$/CD62L$^{-}$; Tcm: central-memory T cells: CD45RA$^{-}$/CD45RO$^{+}$/CD62L$^{+}$; naïve: CD45RA$^{+}$/CD45RO$^{-}$/CD62L$^{+}$. **(B)** Viability of cell product during *ex vivo* manufacturing. Data are shown with color coding by patient (left), by whether starting cell population underwent CD14/CD25 depletion (middle), and by disease indication (right).

**Supplementary Figure S2**

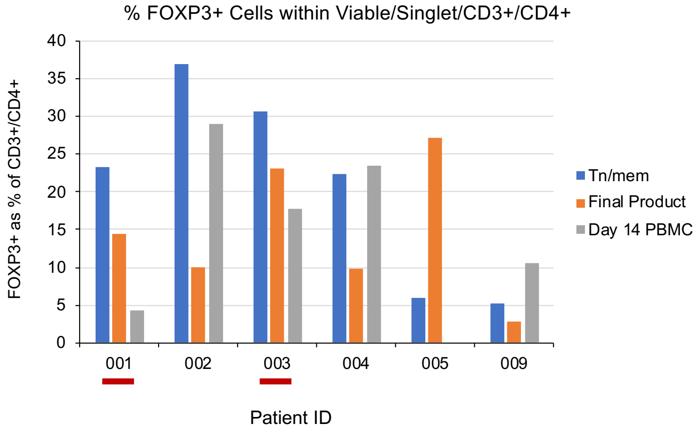


**Supplementary Fig. S2. CD14/CD20 depletion did not significantly alter regulatory T (Treg) cell content in CART19/20 cell product and in patient peripheral blood post infusion.** Intracellular staining of starting (unactivated) $T_{N/MEM}$ population, CART19/20 final product, and patient PBMC collected 14 days post CART19/20 infusion. $FOXP3^{+}$ frequency is shown as a percentage of $CD3^{+}CD4^{+}$ T cells.

### Supplementary Figure S3

**A**

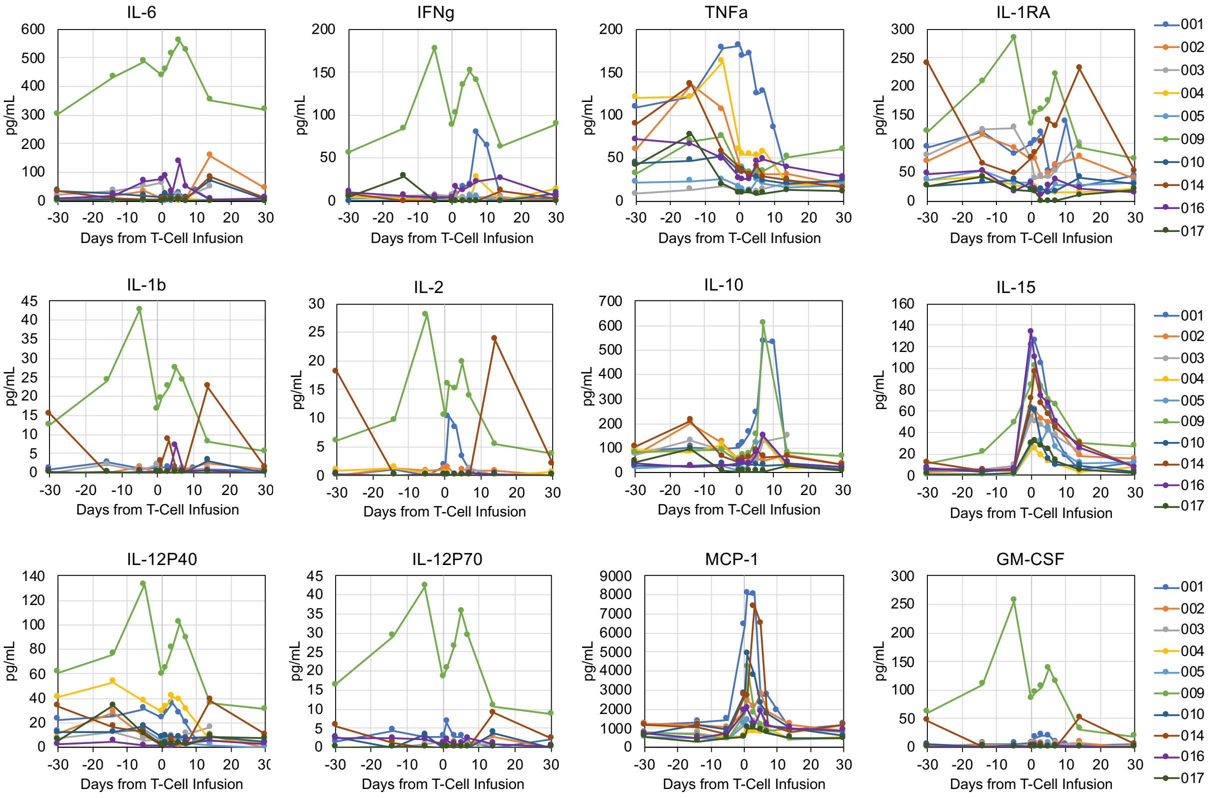


**B**

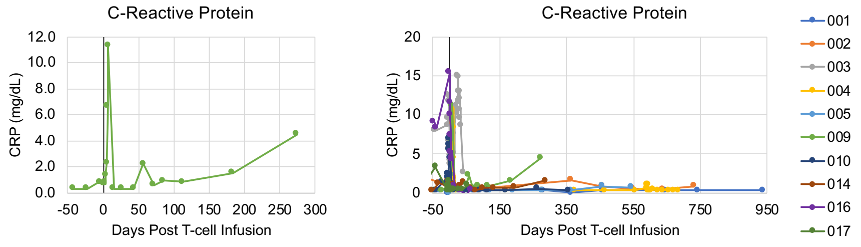


**C**

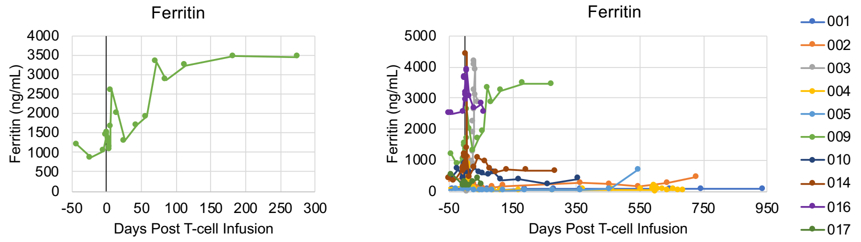


**Supplementary Fig. S3. Patient 009 exhibited elevated cytokine, C-reactive protein (CRP), and ferritin levels prior to and after CART19/20 cell infusion. (A)** Serum levels of various cytokines measured by Luminex multiplex assay. **(B)** CRP levels for Patient 009 (left) and all patients (right). **(C)** Ferritin levels for Patient 009 (left) and all patients (right).

### Supplementary Figure S4

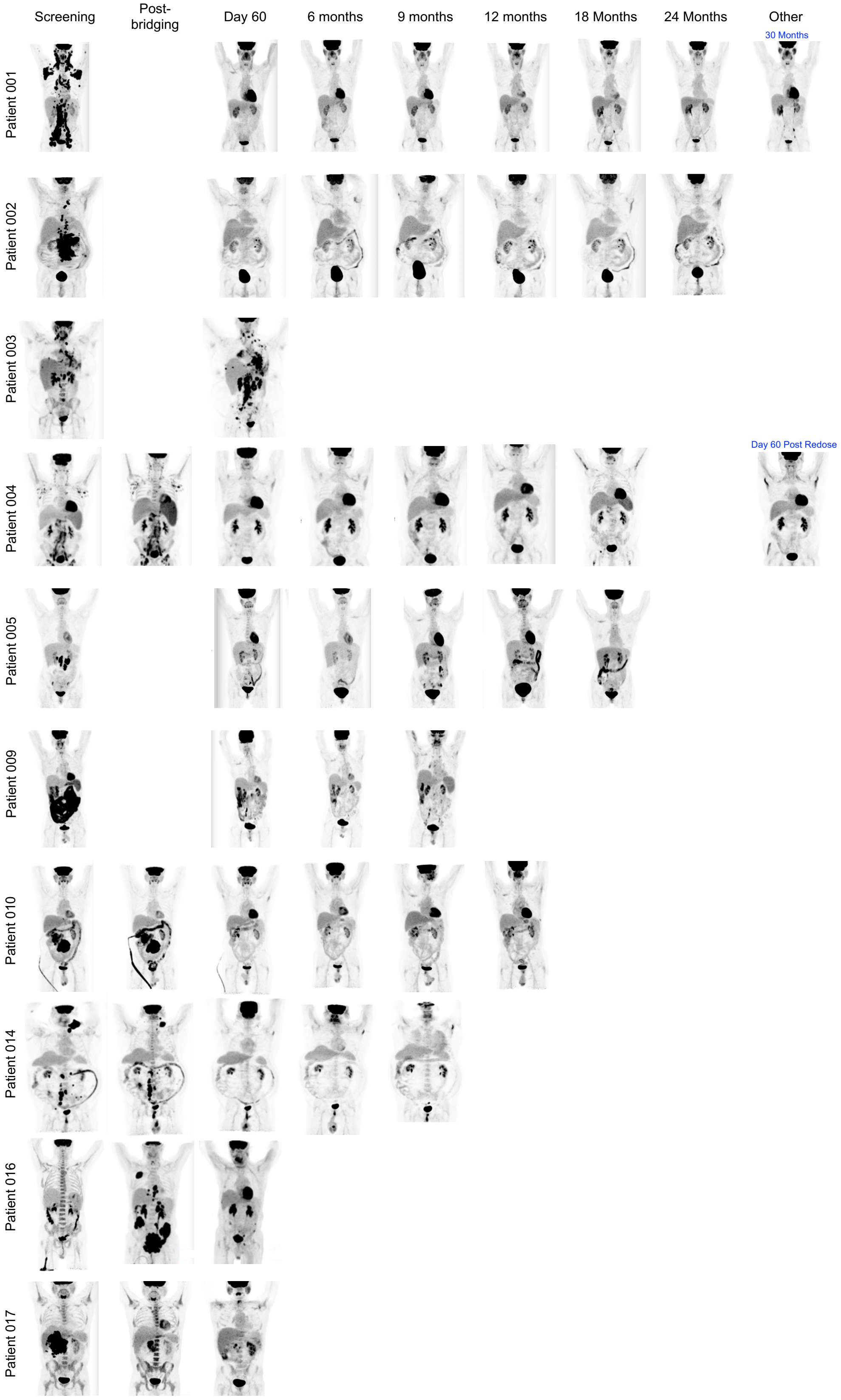

**Supplementary Fig. S4. PET scans taken for all patients treated with CART19/20 cell therapy.**

**Supplementary Figure S5**

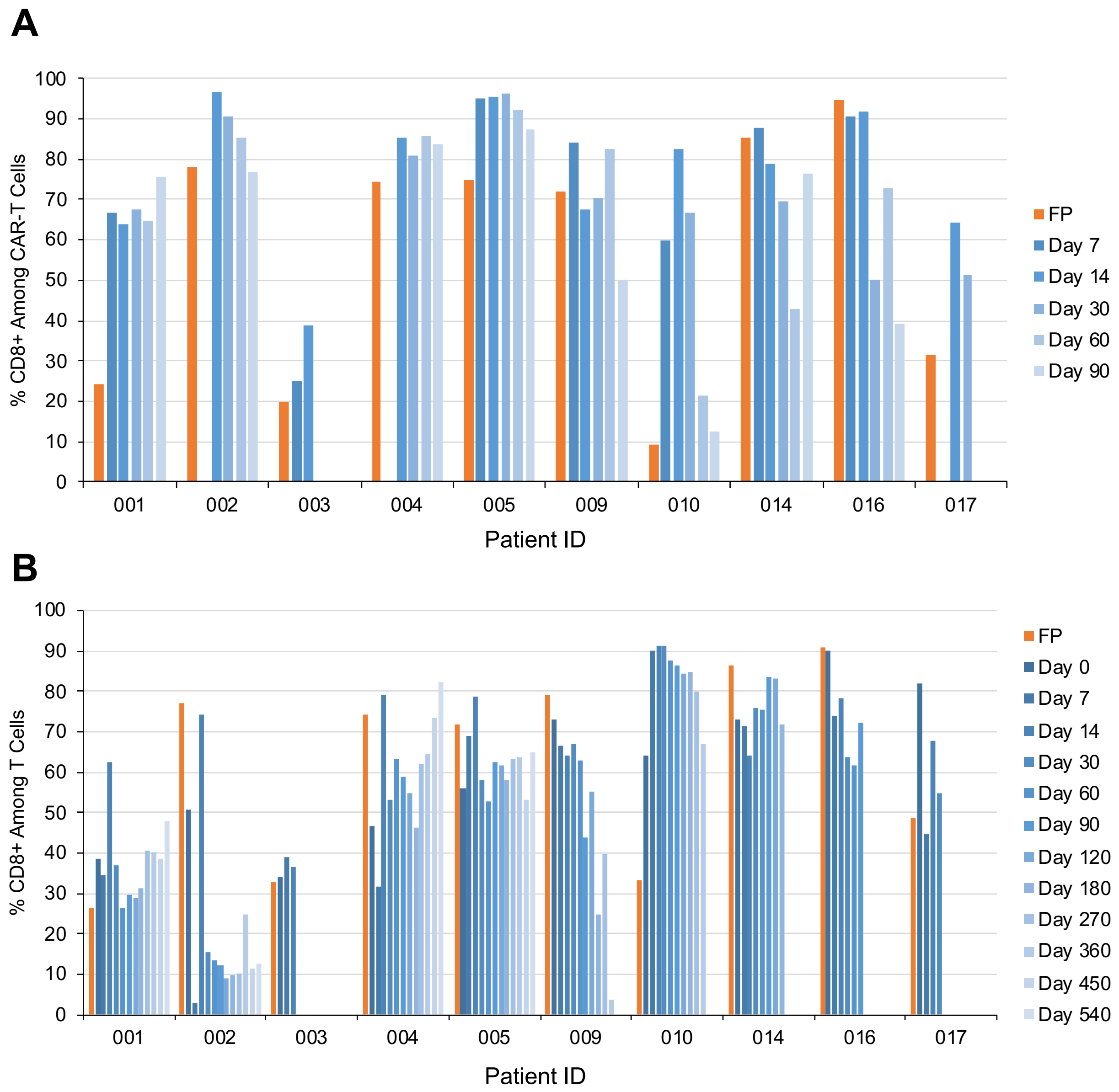


**Supplementary Fig. S5. T cell populations become CD8-dominant post CART19/20 cell infusion.** % $CD8^+$ among **(A)** CAR-expressing T cells and **(B)** all $CD3^+$ T cells was quantified by flow cytometry. Data are shown for CAR-T cells only up to day 90 post infusion as values post day 90 become unreliable due to low $CAR^+$ cell count detected by flow. FP: final product (i.e., cryopreserved CART19/20 cells).
